# A longitudinal cohort study comparing clinical trials registered on ClinicalTrials.gov that stopped during the first three years of the SARS-CoV-2 pandemic with trials that stopped in the three years prior

**DOI:** 10.64898/2026.05.20.26353581

**Authors:** Benjamin Gregory Carlisle, Nora Hutchinson, Hannah Moyer

**Affiliations:** STREAM Research Group, Department of Equity, Ethics and Policy, School of Population and Global Health, McGill University, Montréal Canada; Division of Hopsital Medicine, Department of Medicine, University of California, San Francisco; Temple University

**Keywords:** COVID-19, clinical trials, registry analysis, enrollment disruption, telemedicine, protocol integrity

## Abstract

**Background:** The global SARS-CoV-2 pandemic disrupted healthcare systems worldwide, raising concerns about its impact on clinical research. Early reports suggested reductions in participant enrollment, interruptions to ongoing trials, and challenges to protocol adherence, yet the magnitude and duration of these operational disruptions remain unclear.

**Methods:** We conducted a registry-based analysis comparing clinical trials during the COVID-19 pandemic (December 2019 to November 2022) with a matched pre-pandemic cohort (December 2016 to November 2019). Studies were included if they reported any modifications to trial status, enrollment, or protocols during the study periods. Key variables included trial stoppage, enrollment changes, and adoption of remote or hybrid procedures.

**Results:** The global SARS-CoV-2 pandemic resulted in widespread disruptions to trial operations with 13,323 clinical trials terminated, suspended or withdrawn over the course of the pandemic, a 38% increase compared to the 9,665 trials that stopped in the 3 years prior to the pandemic. Registries indicated a sharp decline in new participant enrollment across geographic regions and therapeutic areas, with partial recovery in later months. Review findings highlighted barriers including patient inaccessibility, staff redeployment, and supply chain interruptions.

**Conclusions:** The pandemic caused system-wide operational shocks that compromised trial timelines and may have downstream methodological consequences. Recovery in enrollment does not imply restoration of pre-pandemic protocol fidelity or outcome ascertainment. Standardized reporting of disruptions, proactive contingency planning, and resilient trial designs are needed to maintain data integrity during large-scale disruptions and to support reliable evidence generation.

**What is new:**

- COVID-19 caused widespread, measurable disruptions to ongoing clinical trials globally, including pauses in enrollment, randomization, and follow-up.
- Recovery of trial activity was uneven across regions and therapeutic areas, with some trials still experiencing delayed recruitment and operational challenges months into the pandemic.
- Standardized reporting of trial disruptions and flexible, resilient trial designs are needed to maintain reliable evidence generation during future large-scale health emergencies.

**Plain language summary:** The COVID-19 pandemic affected the way clinical trials were conducted around the world. Many studies had to stop enrolling new participants, delay treatments, or change how patients were monitored. These interruptions were caused by travel restrictions, social distancing, and limited access to hospitals and research staff. Some studies used phone calls or video visits to continue collecting information, but these changes may have affected the quality of the results. Even as enrollment recovered in some areas, many trials still faced delays and challenges. Understanding these disruptions can help researchers plan better for future emergencies, ensuring that clinical trials remain safe, reliable, and useful for developing new treatments.

## Introduction

The global SARS-CoV-2 pandemic with cases first reported in December 2019 marked worldwide changes in research policy and priority-setting, as well as a change in the feasibility of conducting clinical research in the context of globally expanding public health measures such as curfews, lock-downs and travel bans. These changes re-allocated resources and researcher attention toward clinical trials related to SARS-CoV-2 treatment and prevention and also made recruitment for ongoing trials more challenging or impossible to achieve.

Challenges to successful completion of clinical trials are not unique to the context of a global pandemic. In any given year, many trials are launched and then closed due to unsuccessful accrual, safety concerns, futility, unexpectedly high efficacy, feasibility issues, new information that was not known when the trial was launched, technical or administrative reasons, or re-allocation of resources due to a change in research priorities.^1,2^ Estimates of trial termination put the rate between 12 and 31%.^3,4^

When external events like a global pandemic force the closure of a clinical trial, the possibility that the trial will produce usable medical information is diminished. In addition, the risks and burdens that early enrollees in trials take on may not be redeemed if a trial is stopped before it reaches conclusions. The SARS-CoV-2 pandemic provides a window into the characteristics of trials that were closed, allowing us to take stock of the impact of the pandemic on the world of clinical trials, as well as providing some indication of the kinds of clinical trials that are more likely to be impacted by future external emergencies.

Much is already known about the impact of the COVID-19 pandemic on clinical trials, but existing studies often rely on limited datasets, descriptive analyses, or early observations that fail to capture the full scope and longitudinal patterns of these disruptions. Our study addresses these gaps by analyzing an exhaustive cohort of trials registered on ClinicalTrials.gov over two matched three-year periods, providing a more comprehensive quantitative assessment than prior work. Unlike survey-based approaches such as Chen etal,^5^ our registry-based analysis includes thousands of trials across all disease areas and directly measures operational endpoints including stoppages, enrollment changes, and restart patterns. While descriptive reports like Dorn et al^6^ and registry studies such as Lasch et al^7^ provide valuable insights, our longitudinal design enables direct pre-pandemic comparison and temporal recovery analysis.

Here we report an analysis of an exhaustive data set of all closures of trials registered on ClinicalTrials.gov during the first 3 years of the SARS-CoV-2 pandemic, indexed by the rationale for closure provided by the investigator, and compared to a matching set of clinical trials captured using identical methodology from the 3 years prior to the pandemic.

## Methods

### Primary and secondary outcomes

The primary objective of this project was to estimate the effect of the SARS-CoV-2 pandemic on the conduct of clinical trials registered on ClinicalTrials.gov, by quantifying the number of trial terminations, suspensions and withdrawals in the first three years of the SARS-CoV-2 pandemic (2019-12-01 to 2022-11-30) that were explicitly attributed to the pandemic, as well as the number of patient participants in affected trials. These numbers were compared against the set of all clinical trials that were stopped in the same timeframe (2019-12-01 to 2022-11-30) for reasons that do not cite the pandemic, and the set of all trials that stopped for any reason in the three years prior to the pandemic (2016-12-01 to 2019-11-30).

For our secondary objectives, we focused specifically on interventional trials where at least one patient had been enrolled prior to the trial being terminated, suspended, or withdrawn. This criterion was applied only to our secondary analyses, as we wanted to ensure our assessments of operational disruptions and patient burden were based on trials that had actual participant involvement rather than trials that stopped before any enrollment occurred. Our secondary objectives were 1) to quantify the total number of terminations, suspensions and withdrawals during this time period, regardless of cause, 2) to assess the effect of the SARS-CoV-2 pandemic on subgroups of clinical trials defined by broad disease indication areas, 3) to assess the characteristics of clinical trials that were terminated, suspended or withdrawn because of the pandemic, and 4) to calculate the number of trials that are suspended, terminated or withdrawn and then restarted within one year.

### Data retrieval

To collect the cohort of trials that stopped during the pandmic, all clinical trial registry records from ClinicalTrials.gov that were updated in any way between 2019-12-01 and 2022-11-30 (inclusive) were downloaded for analysis on a rolling basis until 2022-12-20, and only clinical trial records that were “stopped” (overall status changed to “Terminated”, “Suspended” or “Withdrawn”) between 2019-12-01 and 2022-11-30 (inclusive) were saved. If a trial’s overall status was already “Terminated”, “Suspended” or “Withdrawn” prior to 2019-12-01, it would not be included, even if the trial’s “why stopped” field made reference to the SARS-CoV-2 pandemic (e.g. NCT03365921).

If the trial started again (overall status changed from “Terminated”, “Suspended” or “Withdrawn” to any other status) after being stopped according to the definition above by the data cutoff (2022-11-30), the date that the trial restarted and the overall status that it was changed to was also recorded. To allow for at least one year of follow-up after a trial stopped, all trials that stopped during the pandemic were checked again for whether they restarted on 2023-12-20.

The “why stopped” field containing the reason that the trial was stopped, as reported on the first stopped historical version of the clinical trial registry entry on ClinicalTrials.gov, was recorded. The reasons for stopping were manually coded by BGC for whether they indicated explicitly that the trial was stopped due to the SARS-CoV-2 pandemic, including terms such as “Covid-19”, “COVID19”, “Covid”, “NCoV-2019”, “Coronavirus”, “Corona virus”, “Corona”, etc. The complete list of “why stopped” field entries that were included are available from the project repository on the Open Science Framework.^8^ Trials that cite *waning* levels of SARS-CoV-2 infections, etc. as their rationale for stopping were *not* considered to be stopped due to the SARS-CoV-2 pandemic (e.g. NCT04390191).

The “why stopped” field was also manually coded by BGC for whether or not it indicated that the trial was expected to start again, including terms such as “halted temporarily”, “on pause”, “temporarily suspended”, “will resume”, etc.

A matching cohort of clinical trial registry data were downloaded from the same months, three years earlier, from 2016-12-01 to 2019-11-30 (inclusive), to serve as a baseline for comparison.

Other clinical trial registry data were automatically downloaded from ClinicalTrials.gov XML files, such as trial title, summary and detailed description, primary completion date, actual or anticipated enrollment, phase number, study type, patient allocation, intervention model, primary purpose, masking, indications, interventions and sponsors. Trial data were downloaded from ClinicalTrials.gov using an *R* script written for this project. Historical versions of clinical trial records were downloaded using the *R* package cthist.^9^ Broad indication areas were assigned to clinical trials using the TrialFociMapper package in *R* that identifies broad disease indication areas by parsing the MeSH headings that have been attributed to the trial (https://github.com/sama9767/TrialFociMapper/).

### Data integrity

A random sample of 100 trials was triple-coded by BGC, NH and HM and inter-rater agreement among the three raters was calculated by Light’s kappa using the irr *R* package.^10^ As the Light’s kappa was 1 for this random sample, we deemed it unnecessary to perform data extraction in duplicate. The code and ratings are available in the irr folder of the project OSF repository.^8^

### Data availability and dissemination

These data were made openly available online as they were gathered over the course of the pandemic and presented in part in an infographic in Scientific American,^11^ and informed the CONSERVE 2021 Statement.^12^ Here we present the full analysis and data set, available from the project OSF repository.^8^

### Open science statement

The protocol for this project was preregistered with the Open Science Framework before data collection; detailed protocol deviations as well as the data and analysis code for this project are available at the same address.^8^ The two databases, trials that stopped during the first 3 years of the SARS-CoV-2 pandemic, and the comparator of trials that stopped in the 3 years prior, were assembled into an *R* package, ctcovidstop, that is available on Github (https://github.com/bgcarlisle/ctcovidstop/) or OSF.^8^

## Statistics

All statistical tests were two-sided, defining *p* < 0.05 to be statistically significant. Statistical tests are exploratory only, and there was no correction for multiple testing. Analysis of data was performed using *R* v. 4.2.0.^13^

## Results

Over the course of the first 3 years of the SARS-CoV-2 pandemic (2019-12-01 to 2022-11-30, inclusive), 13,323 clinical trials were terminated, suspended or withdrawn. Among the clinical trials that stopped during the pandemic, 3226 (24%) were stopped for reasons that cited the ongoing pandemic. We also identified 9665 clinical trials that stopped in the 3 years prior to the beginning of the pandemic (2016-12-01 to 2019-11-30, inclusive). We estimate that the excess number of clinical trials that were stopped in the first 3 years of the pandemic over the number of stopped clinical trials from the 3 years prior was 3658 clinical trials.

The earliest date on which a clinical trial was stopped, explicitly citing the SARS-CoV-2 pandemic, was on 2020-02-11, when NCT03587103 and NCT03636334 both stopped due to “The outbreak of NCoV-2019 across China.” On average, 84 clinical trials (range 1–311, standard deviation 38) were stopped per week for the first three years of the pandemic (see Figure 1). In the comparator sample of trials that were stopped in the 3 years prior to the beginning of the SARS-CoV-2 pandemic, there was an average of 62 clinical trials (range 14– 106, standard deviation 15) stopped per week (see Figure 2). Starting in the second week of March 2020, there was at least one clinical trial stopped per week, every week, until data cutoff in November 2022. The last week of March 2020 contained the greatest number of clinical trial stoppages due to Covid-19 with 199 trials stopped in that week attributed to the pandemic.

**Figure 1.**
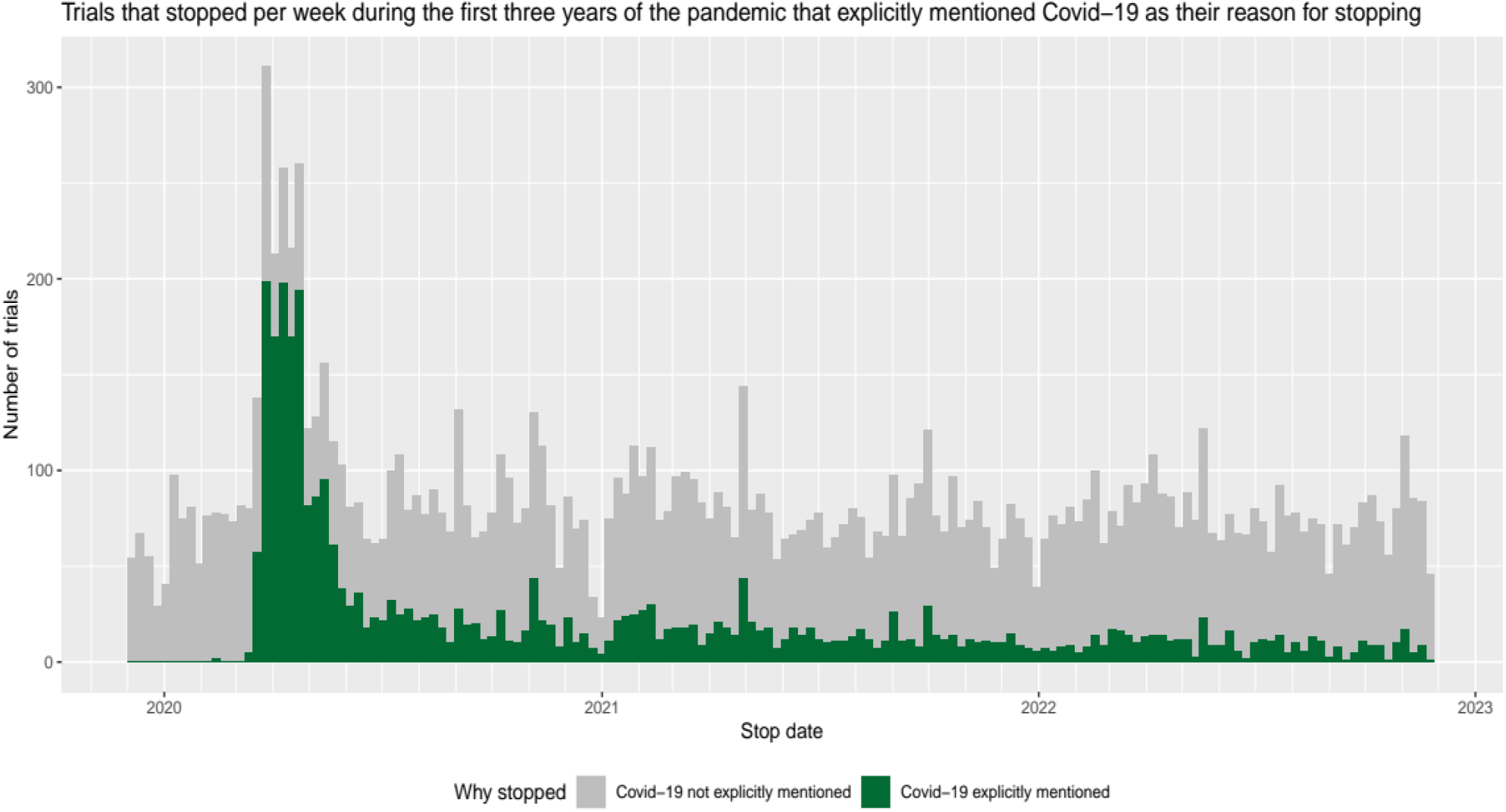
Trials that stopped per week during the first three years of the SARS-CoV-2 pandemic that did not mention the pandemic explicitly (grey) stacked over trials that explicitly mentioned the pandemic as their reason for stopping (green)

**Figure 2.**
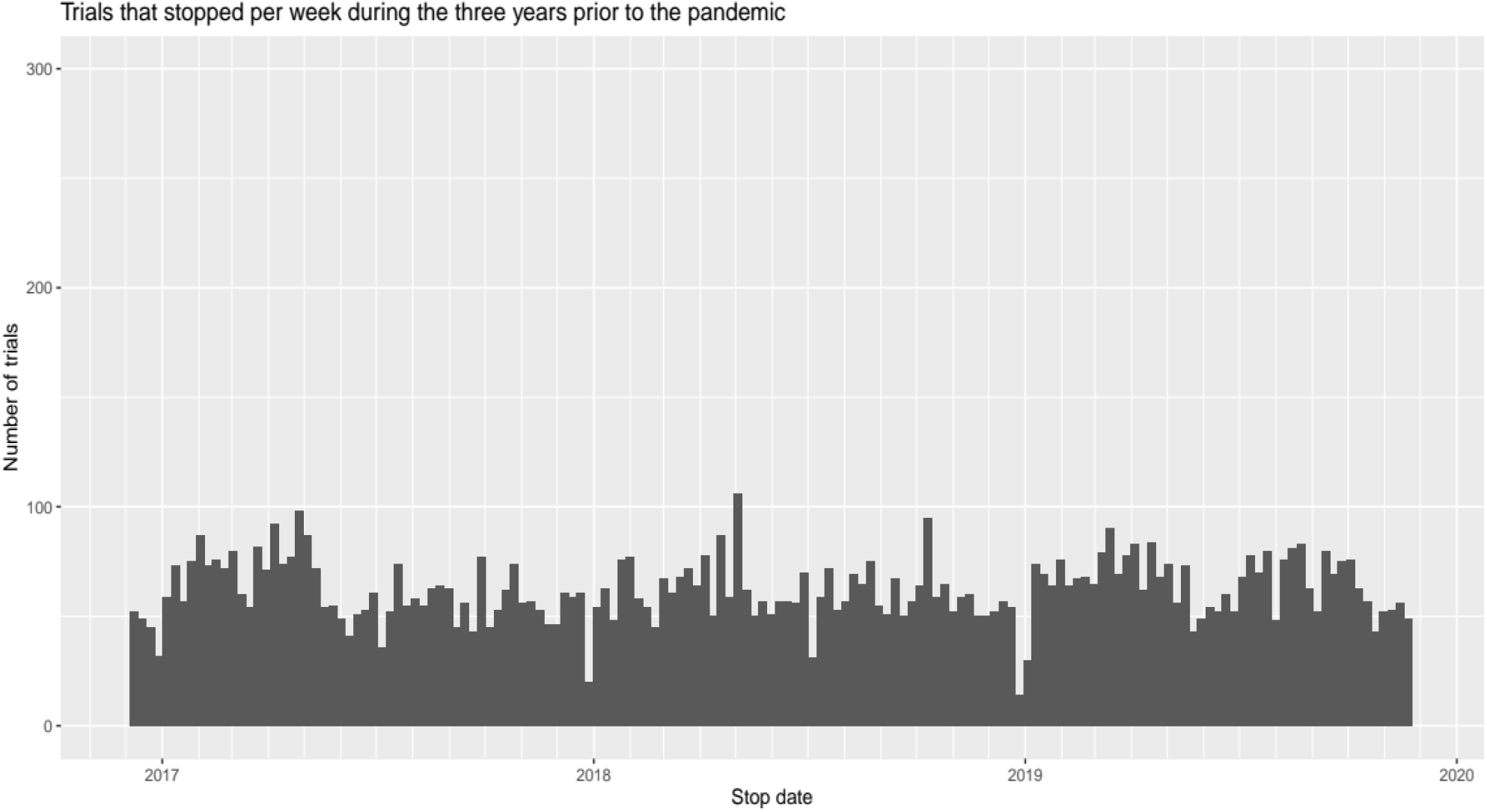
Trials that stopped per week during the three years prior to the pandemic

Among trials that stopped due to the SARS-CoV-2 pandemic, our sample contained 2715 interventional trials and 511 observational trials. Anticipated enrolment was reported for 706 trials, and among the remaining trials with actual enrolment reported, 449 had an enrolment number of zero, leaving 2073 trials with an actual enrolment greater than zero. There were 1782 interventional trials with a non-zero reported actual enrolment. The remaining analyses will focus on this subgroup only (see Table 1).

**Table 1.**
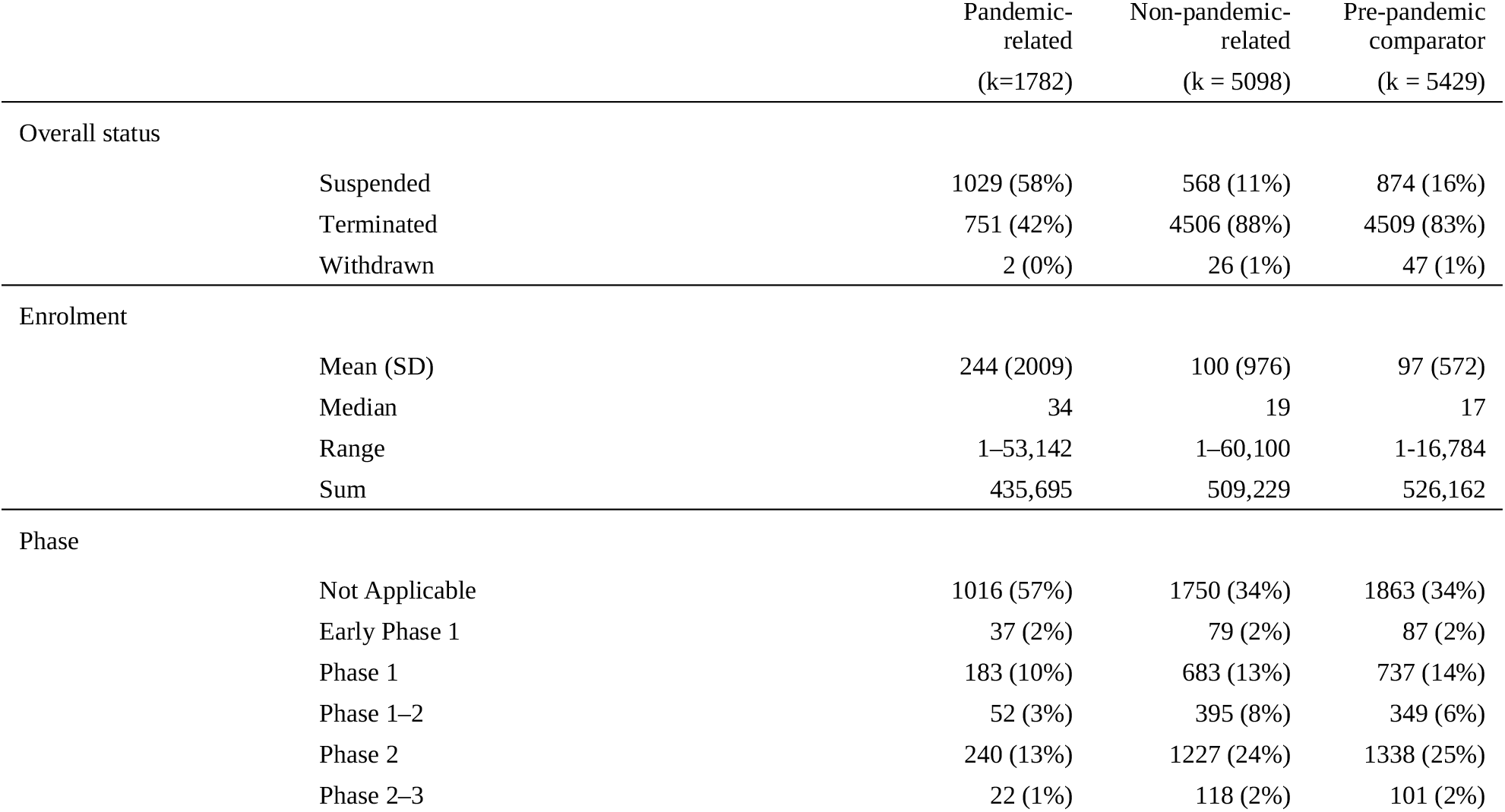
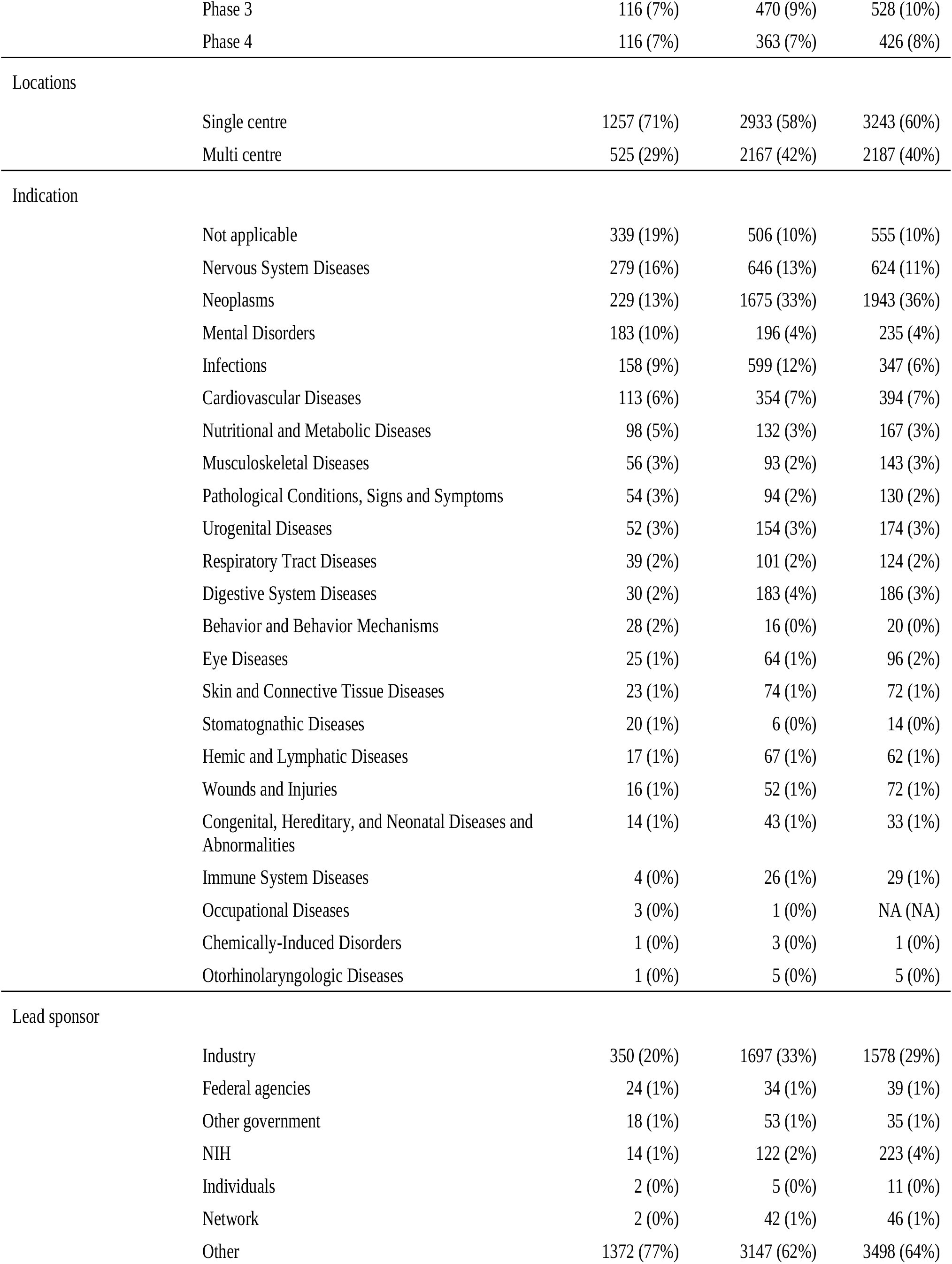
Sample demographics for interventional trials with an actual enrolment of at least one patient among three subgroups: trials that stopped during the first three years of the pandemic for reasons explicitly citing the SARS-CoV-2 pandemic (“Pandemic related”), trials that stopped during the first three years of the pandemic but did not cite the pandemic as their reason for stopping (“Non-pandemic-related”), and trials that stopped during the three years prior to the pandemic (“Pre-pandemic comparator”).

|  |  | Pandemic-related<br>(k=1782) | Non-pandemic-related<br>(k = 5098) | Pre-pandemic<br>comparator<br>(k = 5429) |
| --- | --- | --- | --- | --- |
| Overall status |  |  |  |  |
|  | Suspended | 1029 (58%) | 568 (11%) | 874 (16%) |
|  | Terminated | 751 (42%) | 4506 (88%) | 4509 (83%) |
|  | Withdrawn | 2 (0%) | 26 (1%) | 47 (1%) |
| Enrolment |  |  |  |  |
|  | Mean (SD) | 244 (2009) | 100 (976) | 97 (572) |
|  | Median | 34 | 19 | 17 |
|  | Range | 1–53,142 | 1–60,100 | 1–16,784 |
|  | Sum | 435,695 | 509,229 | 526,162 |
| Phase |  |  |  |  |
|  | Not Applicable | 1016 (57%) | 1750 (34%) | 1863 (34%) |
|  | Early Phase 1 | 37 (2%) | 79 (2%) | 87 (2%) |
|  | Phase 1 | 183 (10%) | 683 (13%) | 737 (14%) |
|  | Phase 1–2 | 52 (3%) | 395 (8%) | 349 (6%) |
|  | Phase 2 | 240 (13%) | 1227 (24%) | 1338 (25%) |
|  | Phase 2–3 | 22 (1%) | 118 (2%) | 101 (2%) |
|  | Phase 3 | 116 (7%) | 470 (9%) | 528 (10%) |
|  | Phase 4 | 116 (7%) | 363 (7%) | 426 (8%) |
| Locations |  |  |  |  |
|  | Single centre | 1257 (71%) | 2933 (58%) | 3243 (60%) |
|  | Multi centre | 525 (29%) | 2167 (42%) | 2187 (40%) |
| Indication |  |  |  |  |
|  | Not applicable | 339 (19%) | 506 (10%) | 555 (10%) |
|  | Nervous System Diseases | 279 (16%) | 646 (13%) | 624 (11%) |
|  | Neoplasms | 229 (13%) | 1675 (33%) | 1943 (36%) |
|  | Mental Disorders | 183 (10%) | 196 (4%) | 235 (4%) |
|  | Infections | 158 (9%) | 599 (12%) | 347 (6%) |
|  | Cardiovascular Diseases | 113 (6%) | 354 (7%) | 394 (7%) |
|  | Nutritional and Metabolic Diseases | 98 (5%) | 132 (3%) | 167 (3%) |
|  | Musculoskeletal Diseases | 56 (3%) | 93 (2%) | 143 (3%) |
|  | Pathological Conditions, Signs and Symptoms | 54 (3%) | 94 (2%) | 130 (2%) |
|  | Urogenital Diseases | 52 (3%) | 154 (3%) | 174 (3%) |
|  | Respiratory Tract Diseases | 39 (2%) | 101 (2%) | 124 (2%) |
|  | Digestive System Diseases | 30 (2%) | 183 (4%) | 186 (3%) |
|  | Behavior and Behavior Mechanisms | 28 (2%) | 16 (0%) | 20 (0%) |
|  | Eye Diseases | 25 (1%) | 64 (1%) | 96 (2%) |
|  | Skin and Connective Tissue Diseases | 23 (1%) | 74 (1%) | 72 (1%) |
|  | Stomatognathic Diseases | 20 (1%) | 6 (0%) | 14 (0%) |
|  | Hemic and Lymphatic Diseases | 17 (1%) | 67 (1%) | 62 (1%) |
|  | Wounds and Injuries | 16 (1%) | 52 (1%) | 72 (1%) |
|  | Congenital, Hereditary, and Neonatal Diseases and Abnormalities | 14 (1%) | 43 (1%) | 33 (1%) |
|  | Immune System Diseases | 4 (0%) | 26 (1%) | 29 (1%) |
|  | Occupational Diseases | 3 (0%) | 1 (0%) | NA (NA) |
|  | Chemically-Induced Disorders | 1 (0%) | 3 (0%) | 1 (0%) |
|  | Otorhinolaryngologic Diseases | 1 (0%) | 5 (0%) | 5 (0%) |
| Lead sponsor |  |  |  |  |
|  | Industry | 350 (20%) | 1697 (33%) | 1578 (29%) |
|  | Federal agencies | 24 (1%) | 34 (1%) | 39 (1%) |
|  | Other government | 18 (1%) | 53 (1%) | 35 (1%) |
|  | NIH | 14 (1%) | 122 (2%) | 223 (4%) |
|  | Individuals | 2 (0%) | 5 (0%) | 11 (0%) |
|  | Network | 2 (0%) | 42 (1%) | 46 (1%) |
|  | Other | 1372 (77%) | 3147 (62%) | 3498 (64%) |

### Patients affected

There was a combined actual enrolment of 435,695 patients in interventional trials that stopped due to the pandemic in the first three years (see Table 1). There was a median of 34 patients per trial, as well as a high variability in trial enrolment (range 1–53,142; mean 245; standard deviation 2010, see Figure 3). Figure 4 shows the cumulative enrolment of patients over time in trials that stopped due to the pandemic. Nearly half of the patients enrolled in trials that stopped due to the pandemic were in trials that stopped in the months of March and April 2020 (45%, 195,891).

**Figure 3.**
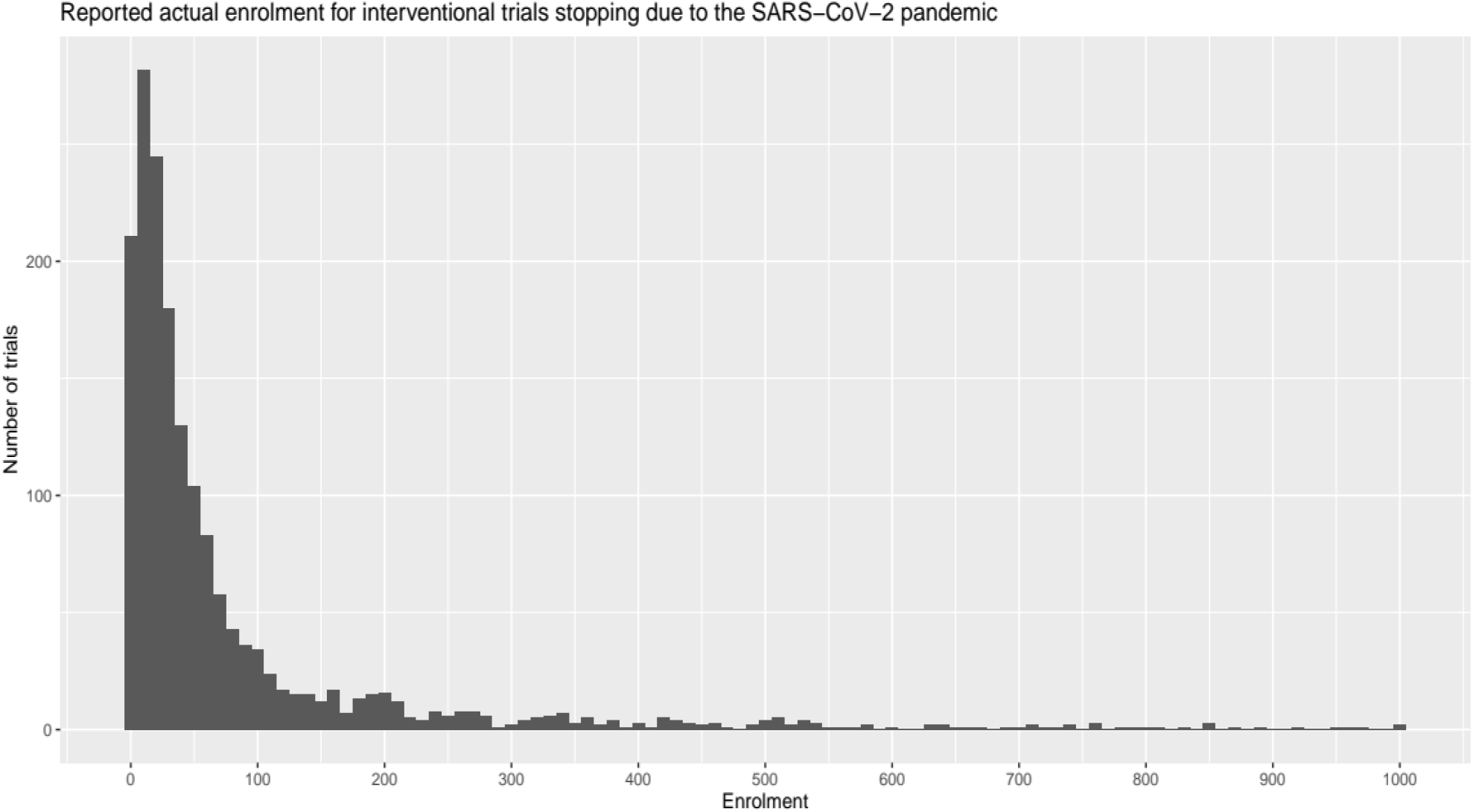
Reported actual enrolment for interventional trials stopping due to the SARS-CoV-2 pandemic. For legibility, this histogram only includes trials with enrolment less than or equal to 1000 patients, omitting 45 trials with enrolment greater than 1000.

**Figure 4.**
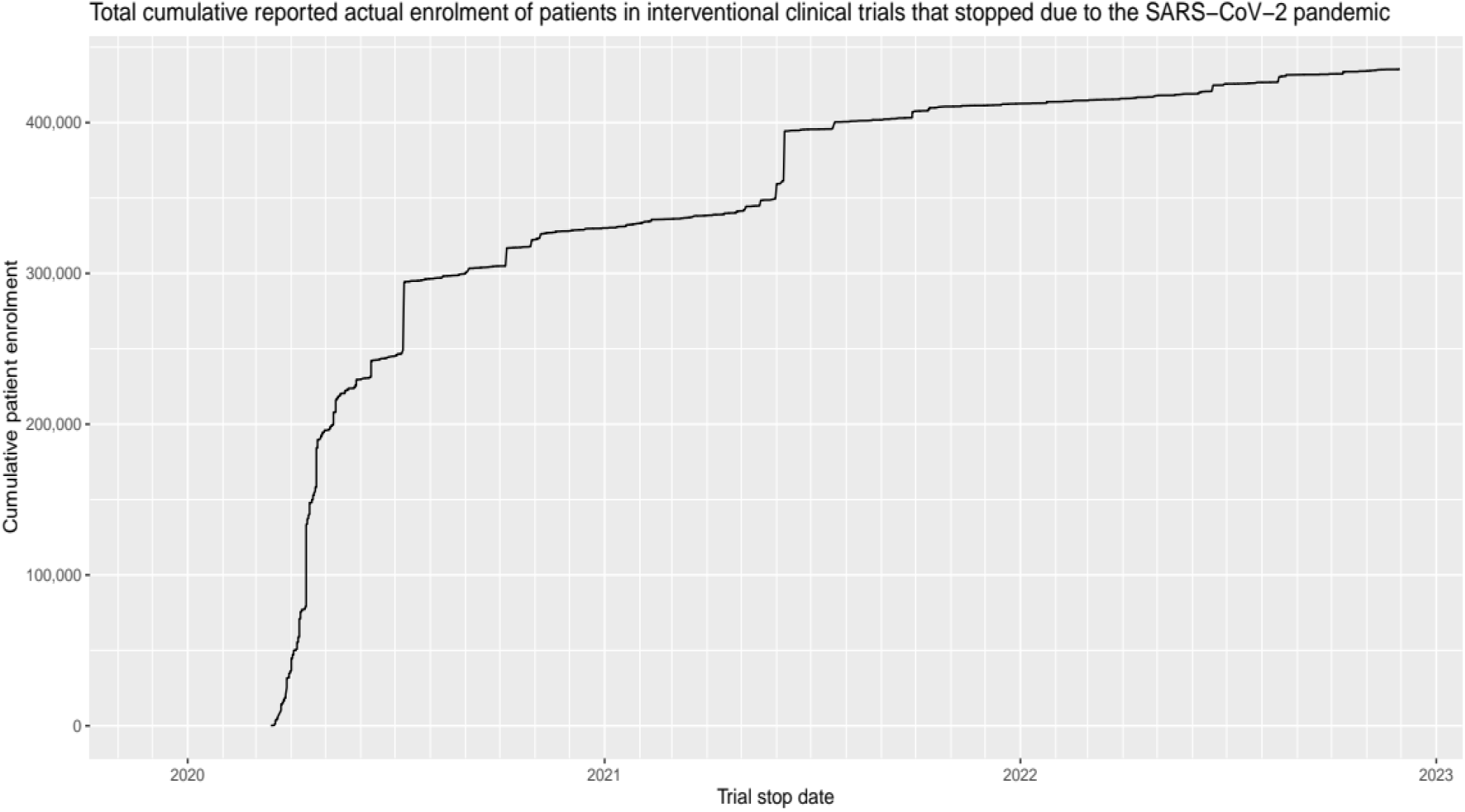
Total cumulative reported actual enrolment of patients in interventional clinical trials that stopped due to the SARS-CoV-2 pandemic

### Trials that started again after stopping

Among interventional clinical trials enrolling at least one patient that stopped in the three years prior to the pandemic, 559 of 5429 (10%) started again within one year. Among interventional clinical trials enrolling at least one patient that stopped during the pandemic without citing the pandemic as the reason the trial stopped, 317 of 5098 (6%) started again within one year. Among trials that stopped for reasons citing the pandemic, 685 of 1782 (38%) started again. Among the 450 interventional trials with at least one enrolled patient that stopped due to the pandemic and mentioned that they expected to start again, 328 (73%) started again within one year.

In clinical trial indications we identified among trials that stopped citing the pandemic as a reason, there was a wide range of rates of trials starting again, from 0% in Otorhinolaryngologic Diseases, to 68% in Neoplasms (see Table 2).

**Table 2.** Proportions of clinical trials that started again within 1 year among three subgroups: trials that stopped during the first three years of the pandemic for reasons explicitly citing the SARS-CoV-2 pandemic (“Pandemic related”), trials that stopped during the first three years of the pandemic but did not cite the pandemic as their reason for stopping (“Non-pandemic-related”), and trials that stopped during the three years prior to the pandemic (“Pre-pandemic comparator”), stratified by indication.

| Indication | Restarted within 1 year<br>(pandemic-related) | Restarted within 1 year<br>(non-pandemic-related) | Restarted within 1 year<br>(pre-pandemic comparator) |
| --- | --- | --- | --- |
| Neoplasms | 155/229 (68%) | 173/1675 (10%) | 337/1943 (17%) |
| Congenital, Hereditary, and Neonatal Diseases and Abnormalities | 9/14 (64%) | 1/43 (2%) | 3/33 (9%) |
| Skin and Connective Tissue Diseases | 12/23 (52%) | 3/74 (4%) | 7/72 (10%) |
| Respiratory Tract Diseases | 20/39 (51%) | 4/101 (4%) | 7/124 (6%) |
| Hemic and Lymphatic Diseases | 8/17 (47%) | 6/67 (9%) | 11/62 (18%) |
| Digestive System Diseases | 14/30 (47%) | 3/183 (2%) | 9/186 (5%) |
| Eye Diseases | 10/25 (40%) | 3/64 (5%) | 8/96 (8%) |
| Stomatognathic Diseases | 8/20 (40%) | 0/6 (0%) | 0/14 (0%) |
| Infections | 63/158 (40%) | 36/599 (6%) | 31/347 (9%) |
| Urogenital Diseases | 20/52 (38%) | 3/154 (2%) | 2/174 (1%) |
| Not applicable | 126/339 (37%) | 29/506 (6%) | 42/555 (8%) |
| Nutritional and Metabolic Diseases | 34/98 (35%) | 5/132 (4%) | 13/167 (8%) |
| Wounds and Injuries | 5/16 (31%) | 0/52 (0%) | 3/72 (4%) |
| Pathological Conditions, Signs and Symptoms | 16/54 (30%) | 6/94 (6%) | 7/130 (5%) |
| Nervous System Diseases | 81/279 (29%) | 27/646 (4%) | 35/624 (6%) |
| Mental Disorders | 53/183 (29%) | 7/196 (4%) | 16/235 (7%) |
| Behavior and Behavior Mechanisms | 8/28 (29%) | 0/16 (0%) | 1/20 (5%) |
| Cardiovascular Diseases | 29/113 (26%) | 7/354 (2%) | 21/394 (5%) |
| Immune System Diseases | 1/4 (25%) | 0/26 (0%) | 4/29 (14%) |
| Musculoskeletal Diseases | 13/56 (23%) | 4/93 (4%) | 2/143 (1%) |
| Chemically-Induced Disorders | 0/1 (0%) | 0/3 (0%) | 0/1 (0%) |
| Occupational Diseases | 0/3 (0%) | 0/1 (0%) | - |
| Otorhinolaryngologic Diseases | 0/1 (0%) | 0/5 (0%) | 0/5 (0%) |
| Endocrine System Diseases | - | 0/10 (0%) | 0/4 (0%) |
| All | 685/1782 (38%) | 317/5098 (6%) | 559/5429 (10%) |

### Indications affected

Per-indication proportions of clinical trials that stopped in the three years prior to the pandemic is similar to the proportions of clinical trials that stopped during the pandemic in trials that did not cite the pandemic as their reason for stopping, with the exception of Infections, which was only 6% in the pre-pandemic cohort and 12% during the pandemic (see Table 1).

There was an elevated proportion of trials of Nervous System Diseases among those that stopped due to the pandemic compared to the three years prior (16% vs 11%, *p* < 0.0001), and a decreased proportion of clinical trials of Neoplasms that stopped due to the pandemic compared to the three years prior (13% vs 36%, *p* < 0.0001).

## Discussion

The global SARS-CoV-2 pandemic increased the number of clinical trial closures in the first three years by about one third (5429 to 6880). Trials closed due to Covid a combined enrolment of 435,695 patients. Nearly half of these trials started again.

The characteristics of trials that stopped in the three years prior to the pandemic are nearly identical to the set of trials that stopped during the pandemic but without citing the pandemic as the reason for stopping. The number of trials, their mean, median, total enrolment, distribution among phases and indications (except Infections) are all within 10% of each other. Furthermore, the number of trials reported as stopped due to the pandemic, based on the stated reasons in the ClinicalTrials.gov records, is very similar to the estimate of excess trials stopped during the pandemic as compared to the pre-pandemic period (3,226 vs. 3,658). This suggests that the reasons cited on ClinicalTrials.gov records for why a trial is stopped are accurate—a trial that says it was stopped due to the pandemic was, very likely stopped due to the pandemic. Even more compelling, the rate at which trials start again increases to 84% among trials where the reason why it stopped seems to indicate that it will start again.

The observed increases in trial stoppages and reductions in enrolment likely reflect multiple concurrent mechanisms operating at the patient, site, and sponsor levels. Public health restrictions and infection risk reduced patient willingness and ability to attend in-person study visits, while research personnel were frequently redeployed to clinical duties or constrained by institutional shutdowns. Sponsors also implemented precautionary pauses or protocol modifications to mitigate uncertainty regarding data completeness and safety monitoring. These layered disruptions suggest that the pandemic acted not as a single operational interruption but as a system-wide shock affecting nearly all components of trial conduct simultaneously.

Trials investigating Nervous System Diseases were more susceptible to being stopped during the SARS-CoV-2 pandemic. This may be because the populations in which these diseases are studied tend to be more at risk for serious injury from SARS-CoV-2, or because of other properties of the trials themselves.^14^ Trials investigating treatments for Neoplasms tended to be less prone to stoppage during the SARS-CoV-2 pandemic. This may be because many cancer therapies are life-saving or -extending, and in many cases, treatment can be administered in out-patient settings.

While stoppages were trailing off toward the end of the data collection period for this study, there was at least one trial that was stopped due to the pandemic every week even until the week of 2022-11-30, suggesting either a long tail of after-effects that was still causing trials to close, or perhaps simple administrative lag, in which trials that had effectively stopped months earlier were not updated on ClinicalTrials.gov.

Although many trials resumed activity following early-pandemic disruption, restart does not necessarily imply restoration of pre-pandemic protocol fidelity or data completeness. Missed visits, altered assessment schedules, and increased reliance on remote procedures may introduce informative missingness or measurement variability that persists beyond the period of operational interruption. Consequently, the apparent recovery of enrolment and trial status metrics may underestimate the downstream methodological consequences of disruption, particularly for time-to-event endpoints and outcomes requiring standardized in-person assessment.

The pandemic also accelerated adoption of decentralized and hybrid trial procedures, including remote monitoring, telemedicine visits, and flexible consent processes. While many of these adaptations were introduced as temporary mitigation strategies, their persistence in post-pandemic trial conduct suggests the possibility of a durable structural shift in clinical research operations. Whether these changes ultimately improve efficiency without compromising data quality remains an important open question.

These findings in this study also have implications for trial reporting standards. Improved structured reporting of operational disruptions, protocol modifications, and missing outcome assessments would enhance the ability of registry-based research to quantify the methodological consequences of future system-level shocks.

The SARS-CoV-2 pandemic disrupted clinical trials through multiple mechanisms. Institutional restrictions prioritized clinical care and COVID-19-related research, delaying or halting non-essential trials (consistent with Chen et al^5^ and Dorn et al^6^). Economic challenges, including funding reallocation toward pandemic-related studies, further contributed to stoppages. Rigid trial designs reliant on in-person visits struggled to adapt to travel restrictions and limited healthcare access, while global supply chain interruptions affected the availability of investigational products and essential materials (as noted in systematic reviews by Sathian et al^15^). Concerns about patient safety reduced willingness to participate in trial activities, particularly for vulnerable populations. These layered disruptions operated simultaneously, creating a system-wide shock that affected nearly all components of trial conduct. Incorporating contingency planning for large-scale disruptions into trial protocols, developing more flexible and resilient trial designs, and standardizing reporting of operational disruptions (as noted in the CONSERVE 2021 statement^12^) may further improve resilience and maintain data integrity during future emergencies. These findings also have important policy implications for research funding agencies, institutional review boards, and regulatory bodies in preparing for future large-scale health emergencies.

## Limitations

This study should be read in light of the following limitations:

We did not attempt to calculate a ‘denominator’ for the number of trials that stopped during the SARS-CoV-2 pandemic, as it was unclear what a meaningful denominator would be; rather we provided the pre-pandemic arm as a means for evaluating the relative scale of the number of stopped trials during this period.

Due to feasibility, if a trial stopped multiple times during the three-year window of our investigation, we focused solely on the reason provided for the first trial stoppage. A trial that stopped initially for non-pandemic-related reasons, then started again and then stopped for pandemic-related reasons would not be counted as a trial that stopped due to the SARS-CoV-2 pandemic, e.g. NCT04139837.

Even when a trial indicates that it was stopped “due to the pandemic,” it is still difficult to pinpoint the exact reason why it trial was stopped. Trial stoppages in this pandemic may have been influenced by a need to shift resources toward development of medicines and vaccines for SARS-CoV-2, by a desire to shield prospective patient-subjects from exposure to the virus, or by some other practical impediment to completing the trial due to the pandemic context. These results and the database of trial stoppages due to the SARS-CoV-2 pandemic^8^ may play a part in that, by providing a baseline from the SARS-CoV-2 pandemic and gesturing at broad indication areas that are more or less susceptible to trial stoppages. This may aid in planning for future emergencies or pandemic situations.

This study also has limitations inherent to registry-based analyses. Trial status and enrolment fields in ClinicalTrials.gov are updated by sponsors and investigators and may lag real-world operational changes. Definitions of stoppage and restart are not standardized, and registry entries may not fully capture temporary interruptions, protocol deviations, or outcome-level disruptions. As a result, our estimates likely represent conservative measures of operational impact.

The COVID-19 pandemic revealed the vulnerability of clinical trial infrastructure to large-scale disruptions, a concern that has continued relevance in the current research landscape. Recent funding instability affecting NIH-funded trials in 2025 demonstrates that trial fragility extends beyond public health emergencies to include systemic funding challenges. As Patel and colleagues reported, over 200 clinical trials were affected by NIH grant terminations in 2025, creating disruptions similar to those observed during the pandemic, including enrollment halts, protocol modifications, and trial terminations.^16^ This convergence of pandemic-related and funding-related disruptions underscores a broader issue of research environment fragility that threatens trial continuity and data integrity across different types of systemic shocks. Our findings on pandemic-related disruptions therefore have implications not only for future public health emergencies but also for addressing ongoing funding instability in clinical research. Both contexts highlight the need for more resilient trial designs, better contingency planning, and standardized reporting mechanisms to maintain research continuity across various forms of large-scale disruption.

## Conclusion

The COVID-19 pandemic produced substantial and measurable disruptions to global clinical trial operations, including increased stoppages and reduced enrolment relative to pre-pandemic baselines. Although many trials subsequently resumed activity, recovery was heterogeneous across therapeutic areas and did not necessarily imply restoration of pre-pandemic data completeness or protocol fidelity. These findings suggest that pandemic-related operational changes may have downstream implications for effect estimation, statistical power, and regulatory interpretation. Registry-based monitoring provides an important population-level view of these disruptions, but improved reporting standards for operational interruptions and protocol deviations are needed to fully characterize their methodological consequences. Future work should evaluate whether pandemic-era adaptations, particularly decentralized trial procedures, represent temporary accommodations or durable structural shifts in clinical research conduct.

## Data Availability

The protocol for this project was preregistered with the Open Science Framework before data collection; detailed protocol deviations as well as the data and analysis code for this project are available at the same address. The two databases, trials that stopped during the first 3 years of the SARS-CoV-2 pandemic, and the comparator of trials that stopped in the 3 years prior, were assembled into an R package, ctcovidstop, that is available on Github (https://github.com/bgcarlisle/ctcovidstop/) or OSF (https://doi.org/10.17605/OSF.IO/2NQXW).

https://github.com/bgcarlisle/ctcovidstop/

## Acknowledgements

We gratefully acknowledge Carole Federico, Delwen Franzen, Maia Salholz-Hillel, and Peter Grabitz for conversations that led to this project.

## Author declarations

All authors have seen and approved the manuscript.

## Competing interests

The authors declare that we have no competing interests.

## Funding statement

This work received no funding.

## Notes

### Competing Interest Statement

The authors have declared no competing interest.

